# Sex differences in the associations between risk for late-life AD, protective lifestyle factors and cognition in mid-life

**DOI:** 10.1101/2023.01.09.23284340

**Authors:** Qing Qi, Feng Deng, Karen Ritchie, Graciela Muniz-Terrera, Ivan Koychev, Paresh Malhotra, John T. O’Brien, Craig W. Ritchie, Brian Lawlor, Lorina Naci

## Abstract

It is now acknowledged that Alzheimer’s Disease (AD) processes are present decades before the onset of clinical symptoms, but whether lifestyle activities can protect against these early AD processes in mid-life remains poorly understood. Furthermore, the impact of sex as a biological variable on associations between dementia risk, protective lifestyle activities and cognition is unknown. In this study, we aimed to replicate findings from our two recent studies [Deng et al. (2022) and Heneghan et al. (2022)] on the contribution of mid-life modifiable activities to cognition in individuals with dementia risk, in a larger independent cohort of the PREVENT–Dementia research program (N = 461 vs N = 208 used previously). Second, we investigated associations between biological sex, dementia risk, protective lifestyle activities and cognitive performance. Participants (40–59 years; N = 461) completed cognitive and clinical assessments cross-sectionally. Mid-life activities were measured with the Lifetime of Experiences Questionnaire. Known risk factors for sporadic late-onset AD (Apolipoprotein E _Ɛ_4 allele status, family history of dementia, and the Cardiovascular Risk Factors Aging and Dementia score [CAIDE]) were investigated. Replicating our key previous findings (Deng et al., 2022 and Heneghan et al., 2022), we found that episodic and relational memory was (a) significantly negatively associated with the CAIDE risk score, (b) positively associated with stimulating lifestyle activities, and (c) that females performed significantly better than males in episodic and relational memory. The key novel finding of this study was that inherited dementia risk (i.e., APOE _Ɛ_4 genotype) modulated the association between sex, lifestyle and cognition. Only for APOE _Ɛ_4+ females, not APOE _Ɛ_4-, higher occupational attainment was associated with better episodic and relational memory. Conversely, only for APOE _Ɛ_4+ males, not APOE _Ɛ_4-, higher occupational attainment was associated with worse episodic and relational memory. These findings suggest that modifiable lifestyle activities offset cognitive decrements due to inherited AD risk in mid-life and support the targeting of modifiable lifestyle activities for the prevention of Alzheimer’s disease. Furthermore, these findings suggest an urgent need for targeted research on female-specific risk factors, to inform personalised strategies for AD prevention and the promotion of female brain health.

## Introduction

Dementia is a growing epidemic that poses significant issues to caregivers, health care systems, and societies throughout the world (Jutkowitz et al., 2017; Sallim et al., 2015). Alzheimer’s disease (AD), which accounts for 60–70% of dementia cases (Fratiglioni et al., 2000; Mayeux and Stern, 2012), two-thirds female (Alzheimer’s Association, 2017), is characterized by relentless neurodegeneration and accelerated cognitive decline in the years following presentation of clinical symptoms. There is now consensus that neurodegenerative disease, including Alzheimer’s disease, is a disease with onset in mid-life that manifests in later life as dementia and related disorders (Ritchie et al., 2015; Jack Jr et al., 2013). Evidence suggests that up to 40% of future incidence may be prevented by addressing 12 lifestyle modifiable risk factors, many of which (e.g., hypertension, obesity, and alcohol consumption) become predominant in mid-life (Livingston et al., 2020; Norton et al., 2014). Mid-life, thus, presents a critical and unique window for disease-altering interventions, before the manifestation of substantial brain damage. Mid-life is also critical for female-specific prevention strategies, because menopausal transition is a middle-age phenomenon with knock on effects of endocrinal (Weber et al., 2014), cardiovascular (Yanes and Reckelhoff, 2011), and metabolic (Rahman et al., 2020; Anagnostis et al., 2019) systems that significantly impact brain health (Udeh-Momoh and Watermeyer, 2021; Mosconi et al., 2018). However, very little is known on the moderating effect of biological sex on the interplay between risk for late-life AD and protective lifestyle factors on brain health in mid-life.

Biological sex is a key modulator of AD risk (Mazure and Swendsen, 2016; Riedel et al., 2016) that has so far not been adequately considered in studies of AD (Luckey et al., 2021). Not only do females account for over two-thirds of AD cases (Mielke, 2018; Hebert et al., 2013), but they also make up approximately two-thirds of caregivers for individuals with dementia (Alzheimer’s Association, 2021), and thus, are more likely than men to be exposed to negative influences that contribute to AD neuropathology, including increased stress (Ma et al., 2018), caregiver burden (Xiong et al., 2020) and mental health problems (Erol et al., 2015). Little is known, however, on the effect of sex on intervention strategies for optimizing brain health in mid-life.

Studies on preclinical AD have used risk stratification approaches to investigate early, preclinical changes. Key risk factors that have been investigated include Apolipoprotein E (APOE) _Ɛ_4 genotype (Liu et al., 2013; Donix et al., 2012), the main genetic risk factor for sporadic late onset AD in the Indo-European population, and family history of dementia (FHD) (Scarabino et al., 2016; Donix et al., 2012; Berti et al., 2011). Lifestyle also presents several risk factors that have been incorporated in lifestyle-based dementia risk scores (Deckers et al., 2015; Barnes et al., 2014; Kivipelto et al., 2006). Amongst them, the Cardiovascular Risk Factors Aging and Dementia (CAIDE) score, was optimized for middle-aged populations (Kivipelto et al., 2006) and has been validated in a large US population followed longitudinally over 40 years (Exalto et al., 2014).

The PREVENT–Dementia program (Ritchie & Ritchie, 2012) initiated in 2013, is a prospective international study of the cognitively healthy middle-aged children of persons with dementia, designed to seek out clinical and biological changes, which may subsequently be used as short-term outcome measures for mid-life secondary preventions. The program is carried out in four universities in the UK – Imperial College London, the University of Edinburgh, the University of Cambridge, the University of Oxford – and Trinity College Dublin, in Ireland. Studies of this and other similar cohorts of cognitively healthy middle-aged individuals have revealed these three risk factors – APOE _Ɛ_4 genotype, FHD, and CAIDE score – impact cognition and brain health in mid-life. APOE _Ɛ_4 carriership has been associated with impaired attention (Zimmermann et al., 2019), better form perception (Ritchie et al., 2017), better verbal, spatial and relational memory (Deng et al., 2022), decreased volume in the hippocampal molecular layer (Dounavi et al., 2022; Dounavi et al., 2020), and reduced cortical thickness in the entorhinal cortex (McKiernan et al., 2020). FHD has been associated with worse visual working memory (Ritchie et al., 2017), worse visuospatial memory and executive function (Debette et al., 2009), cerebral hyperperfusion (McKiernan et al., 2020), hippocampal volume loss (McKeever et al., 2020), and impaired white matter integrity (Bendlin et al., 2010). Higher CAIDE scores have been associated with impairments in visual recognition (Ritchie et al., 2017), allocentric spatial processing ability (Ritchie et al., 2018), verbal, visuospatial functions and short-term memory (Deng et al., 2022), decreased grey matter volume in the temporal, occipital, and fusiform cortex (Liu et al., 2021), reduced cortical thickness in the temporal cortex (Dounavi et al., 2022; Gourley et al., 2020), and brain volume loss (O’Brien et al., 2020). Taken together, these studies have established that risk for late-life AD, including risk that incorporates lifestyle factors, has a significant impact on the brain health and cognition of middle-aged individuals who are presently cognitively healthy.

Lifestyle has a multipronged impact on cognition and the brain. By contrast to aforementioned risk factors, several lifestyle activities have been associated with preservation of cognitive function in older adults (Chan et al., 2018) and reduced symptom severity in Alzheimer’s disease (Livingston et al., 2020; Dekhtyar et al., 2019; Livingston et al., 2017; Wang et al., 2017), a phenomenon known as “cognitive reserve” (Stern et al., 2020; Stern, 2012). Education, occupational attainment, and physically, socially and intellectually stimulating activities have been identified as key proxies of cognitive reserve. These factors are thought to explain why, in late-life Alzheimer’s disease, the level of cognitive impairment shows substantial variability even when accounting for key pathologies including beta-amyloid (A*β*) and pathological tau (Franzmeier et al., 2020; Jack Jr et al., 2013). Epidemiological evidence strongly suggests that education contributes to cognitive reserve in late life (Richards & Deary, 2005), with its effects set in motion from childhood, and therefore its modifiability in mid-life, while not impossible, is limited. For this reason, there is a renewed interest in the additional contribution of activities undertaken in mid-life, given their modifiability in this period.

Until recently it remained unknown whether lifestyle activities that contribute to cognitive reserve can offset dementia risk, from mid-life, prior to onset of symptoms. A recent study from the PREVENT–Dementia program site at Imperial College London (N = 208) showed for the first time that mid-life stimulating lifestyle activities other than education, can offset the impact of AD risk on cognition in cognitively unimpaired individuals (Heneghan et al., 2022). More frequent engagement in these activities was associated with better episodic and relational memory in at-risk individuals. Heneghan et al. (2022), also found a trend effect of sex on episodic and relational memory, but the effect of biological sex on the relationship between risk factors, lifestyle activities and cognition were not investigated. Since its publication, the cross-sectional study data from the four other sites of the PREVENT– Dementia program has become available (N = 461). The sequential release of these two datasets provided a unique opportunity to validate our previous findings (Heneghan et al., 2022; Deng et al., 2022, N = 208) in a larger sample, and to investigate the associations between sex, dementia risk, protective lifestyle activities and cognitive performance in cognitively healthy middle-aged individuals.

Our objectives in this study were twofold. First, we aimed to replicate findings from two recent studies [Deng et al. (2022) and Heneghan et al. (2022)] on the contribution of mid-life modifiable activities to cognition in individuals with dementia risk, in a larger cohort of the PREVENT–Dementia research program. Second, we investigated associations between biological sex, dementia risk, protective lifestyle activities and cognitive performance. A cohort of cognitively healthy middle-aged individuals was assessed cross-sectionally (N = 461), with the same detailed neuropsychological assessments and research protocols as those published previously (Deng et al., 2022; Heneghan et al., 2022). While the dichotomies of sex and gender are no longer considered to be sharply discrete, in this study ‘sex’ will be defined as an individual’s natal or biological sex. The APOE _Ɛ_4 genotype, FHD, and the CAIDE risk score were used to quantify risk of late-life dementia. Using the same instrument as in these previous studies – the Lifetime of Experiences Questionnaire (LEQ) (Valenzuela & Sachdev, 2007) – we evaluated protective lifestyle activities, yielding two composite factors specific to mid-life, (a) occupational attainment: occupational complexity and managerial responsibility, and (b) physically, socially and intellectually stimulating activities. A data reduction approach was used to obtain the key cognitive dimensions representative of cognitive performance from 13 cognitive measures. We hypothesized that more frequent engagement with stimulating activities in mid-life, other than education, would be associated with stronger cognition in individuals at risk for late-life dementia, and that biological sex would moderate the relationship between risk factors, protective activities and cognition.

## Methods

### Participants

491 participants were recruited in the PREVENT–Dementia program, from four study sites: the University of Cambridge, the University of Oxford, the University of Edinburgh and Trinity College Dublin. The main entry criteria were age between 40 and 59 and absence of dementia or other neurological disorders. The detailed protocol has been described in detail elsewhere (Ritchie & Ritchie, 2012) (Figure 1).

**Figure 1.**
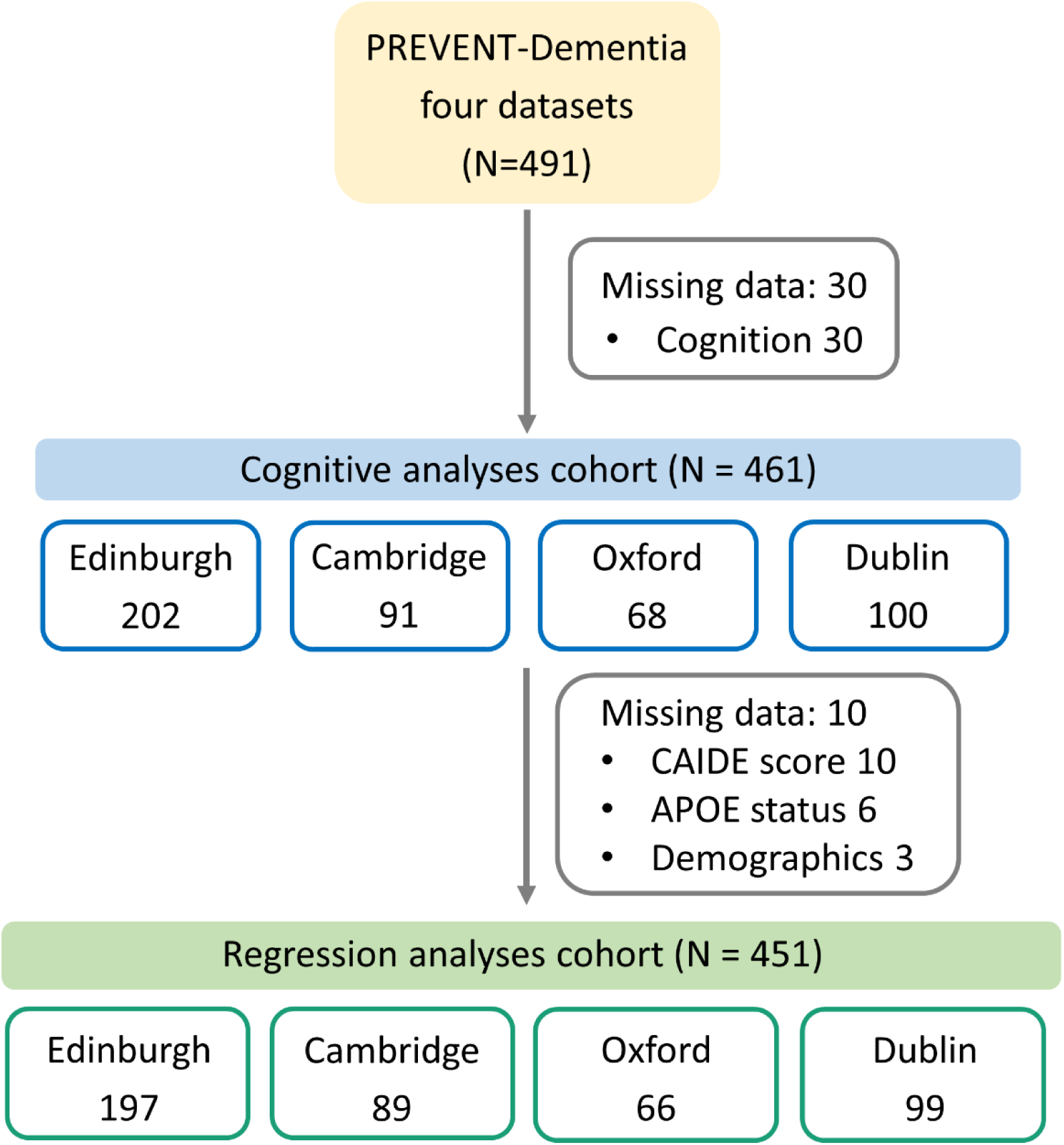
Participant inclusion criteria in the present study for different analyses per site from PREVENT-Dementia program

491 participants (205 male; 286 female) completed cognitive and clinical assessments. For cognitive tests, 30 participants were missing data and the cognitive analyses cohort was N = 461. For clinical assessment, 10 participants were missing information for calculating their CAIDE scores, including the 6 missing APOE status and the 3 missing years of education. Therefore, the regression analyses cohort was N = 451. One participant had missing information in cognitive tests, APOE status, years of education, and LEQ scores, which was regarded as missing data once in the cognitive tests and not counted repeatedly in the subsequent analyses. Please see Figure 1 and Supplementary Table 1 for details of participant inclusion in each analysis.

### Biological sex

Participants self-reported their biological sex in the Brain Injury Screening Questionnaire.

### Risk factors

#### APOE _Ɛ_4 genotyping

The process of APOE _Ɛ_4 allele identification is outlined in detail in Ritchie et al. (2017). In brief, genomic DNA was isolated from blood samples and APOE genotyping was performed. All members of the research and clinical teams were blind to the result of APOE genotyping. In this study, APOE _Ɛ_4 risk is determined by ≥1 APOE _Ɛ_4 allele. 188/491 carried ≥1 APOE _Ɛ_4 allele (See Table 1).

**Table 1.** Demographic specifications of the cohort based on family history of dementia and on APOE genotype

| PREVENT-Dementia program four datasets (n=491) |  |  |  |
| --- | --- | --- | --- |
|  | FHD-<br>(n=234) | FHD+<br>(n=257) | *p*<br>(Mann-Whitney U) |
| Age (y) | 51.0 ± 10.25 | 52.0 ± 7.0 | 0.09 |
| Years of Education | 17.0 ± 4.0<br>(n=230) | 17.0 ± 4.5<br>(n=257) | 0.63 |
| CAIDE (incl. APOE status) | 6.0 ± 5.0<br>(n=228) | 6.0 ± 4.0<br>(n=252) | 0.34 |
| *p* (Chi-Square) |  |  |  |
| Sex (%f) | 54.3% | 61.9% | 0.09 |
| APOE ε4 (% Carriers) | 33.2%<br>(n=232) | 44.0%<br>(n=252) | 0.01 |
|  | APOE ε4-<br>(n=296) | APOE ε4+<br>(n=188) | *p*<br>(Mann-Whitney U) |
| Age (y) | 52.0 ± 9.0 | 51.0 ± 9.0 | 0.10 |
| Years of Education | 17.0 ± 4.0<br>(n=294) | 17.0 ± 5.0<br>(n=187) | 0.64 |
| CAIDE (incl. APOE status) | 6.0 ± 4.0<br>(n=293) | 7.0 ± 4.0<br>(n=187) | <0.001 |
| CAIDE (excl. APOE status) | 6.0 ± 4.0<br>(n=293) | 5.0 ± 4.0<br>(n=187) | 0.24 |
| *p* (Chi-Square) |  |  |  |
| Sex (%f) | 58.1% | 58.0% | 0.98 |

Note: median ± interquartile range (IQR) was reported for continuous variables.
Abbreviations: FHD-, negative family history of dementia; FHD+, positive family
history of dementia; APOE ε4+, Apolipoprotein ε4 genotype positive; APOE ε4-,
Apolipoprotein ε4 genotype negative, CAIDE, Cardiovascular Risk Factors, Aging and Dementia.

#### Family history

FHD was determined by a ‘yes’/‘no’ question during clinical visits, which asked participants whether a parent had a diagnosis of dementia. Participants were asked to include the dementia subtype if known, but answering ‘yes’ alone categorized a participant as FHD+. The answer ‘no’ likely captured both participants with no family history of dementia, and participants for whom FHD was unknown. In summary, participants were defined as FHD+ if at least one parent was diagnosed with dementia. Cases where the FHD was unknown or partially known were not recorded outside of the binary yes/no scoring. 257/491 were FHD+.

#### Cardiovascular Risk Factors, Aging, and Incidence of Dementia (CAIDE) score

CAIDE is a composite scale of estimated future dementia risk based on mid-life cardiovascular measures (Fayosse et al, 2020; Sindi et al., 2015). It takes into consideration the individual’s age, sex, educational attainment, APOE _Ɛ_4 genotype, activity level, BMI, cholesterol and systolic blood pressure (Kivipelto et al., 2006) and is scored on a range of 0 – 18. A higher score indicates greater risk. The CAIDE dementia risk score was calculated for each participant.

### LEQ

The Lifetime of Experiences Questionnaire (LEQ) (Valenzuela & Sachdev, 2007), designed to take a lifespan approach to the measurement of cognitive reserve (Stern et al., 2019; Stern, 2012; Stern, 2009), by measuring engagement in a broad range of lifestyle activities across three stages of life: young adulthood (13-29 years), mid-life (30-64 years) and late life (65 years onwards). Therefore, the LEQ is preferable for looking at a mid-life cohort compared to other scales that capture dementia-specific risk related to modifiable lifestyle factors [e.g., LIBRA (Schiepers et al., 2018)]. The LEQ comprises sub-scores capturing “specific” activities, reflecting the primary activity undertaken in each life stage and “non-specific” activities, reflecting engagement in physical, social and intellectual activities in any stage. For the purpose of this paper, we define mid-life ‘lifestyle’ as all the activities captured by the LEQ (below).

#### Mid-life specific score (Occupational attainment)

The mid-life specific component score centers on occupation and comprises two sub-scores that measure (a) the occupational complexity and (b) the managerial responsibility. For the first occupational sub-score, participants were asked to record their primary occupation in each 5-year interval from age 30 to age at assessment. Each reported occupation was scored on a scale of 0-9, according to the International Standard Classification of Occupations (ISCO 08) guidelines (https://www.ilo.org/public/english/bureau/stat/isco/isco08/). This scale relates to the skill level associated with occupations, where managers score 1, professionals 2, technicians and associate professionals score 3, and so on. Participant scores were inverted and summed. The second sub-score is a measure of the managerial responsibility associated with reported occupations. If participants indicated that they were employed in a managerial capacity, the number of people that they oversaw in four of their reported occupations was documented. Managerial responsibility was scored as follows; 0 people = 8, 1-5 people = 16, 5-10 people = 24 and 10+ people = 32. The highest score is recorded as the managerial responsibility sub-score. The occupational complexity and managerial sub-scores were summed and multiplied by a normalization factor of 0.25. Normalization ensures that the mid-life specific and non-specific scores have comparable mean values (Valenzuela & Sachdev, 2007).

#### Mid-life non-specific score

The non-specific score assesses frequency of engagement in 7 activities, capturing those of a physically, socially and intellectually stimulating nature, scored on a 6-point Likert scale of frequency (never, less than monthly, monthly, fortnightly, weekly, daily). Scores range from 0 – 35, with higher scores reflecting more frequent engagement in such activities. The items included in the scale are socializing with family or friends, practicing a musical instrument, practicing an artistic pastime, engagement in physical activity that is mildly, moderately, or vigorously energetic, reading, practicing a second language and travel. The travel item asks participants if they have visited any of a list of continents between the ages of 30-54. Responses were scored on a 6-point scale as follows: none, 1-2 regions, 3-4 regions, 5 regions, 6 regions, 7 regions.

### Cognitive Assessments

Cognitive function was assessed with the COGNITO neuropsychological battery (Ritchie et al., 2014), designed to examine information processing across a wide range of cognitive functions in adults of all ages and not restricted to those functions usually implicated in dementia detection in the elderly. Tests are administered using a tactile screen to capture information processing time as well as response accuracy and require about 40 minutes to complete. The tests, by order of presentation, are: reaction time; reading; comprehension of phonemes, phrases, and syntax; focused and divided attention in both visual and auditory modalities; visual working memory (visual tracking with auditory interference); the Stroop test; immediate, delayed, and recognition trials for verbal recall (name list); delayed recognition of spatial stimuli (faces); visuospatial associative learning; visuospatial span; form perception; denomination of common objects; spatial reasoning; copying of meaningful and meaningless figures; verbal fluency with semantic and phonetic prompts; immediate recall of a narrative; immediate recall of a description of the relative position of objects; vocabulary; implicit memory (recognition of new and previously learned material).

The COGNITO tests are designed to test several aspects of cognition, including attention (task: visual attention), memory (tasks: narrative recall, description recall, implicit memory, name-face association, working memory), language (tasks: phoneme comprehension, verbal fluency) and visuospatial abilities (task: geometric figure recognition) (Ritchie et al., 2018; Ritchie et al., 2014). Based on previous studies (Ritchie et al., 2018; Ritchie et al., 2014), 11 summary variables from the COGNITO battery capturing the above functions were used here [for a list see Supporting Information (SI)].

Additionally, we used the Visual Short-Term Memory Binding task (VSTMBT) (Parra et al., 2010), a computer-based task that assesses visual short-term memory binding of single features, e.g., complex shape or color combinations, or feature conjunctions, e.g., shape and color combinations. In the single feature condition, participants must identify whether the test stimuli (three random 6-sided polygons) are the “same” as or “different” to the studied stimuli in terms of shape (shape only) or color (color only). In the binding condition, participants are required to correctly identify if both the shape and color of the test stimuli match the studied stimuli. Two summary variables from the VSTMBT were the percentage of correctly recognized items from the two conditions.

### Rotated principal component analysis

Rotated principal component analysis (rPCA), a dimensionality reduction technique, was adopted to cluster the above-mentioned multiple cognitive measures (13 summary variables, see SI and Table S1) into related domains, and reduce the number of multiple comparisons between the cognitive tests. rPCA was conducted in Statistical Package for Social Sciences (SPSS V.27), following steps as: (1) *Preprocessing*. Listwise deletion procedure was used to deal with the missing data. Only subjects who had no missing observations were kept (N = 461). To assess the adequacy of conducting rPCA analysis on this dataset, Kaiser–Meyer–Olkin (KMO) Measure of Sampling Adequacy and Bartlett’s test of the sphericity was used on the dataset. As the KMO value was larger than 0.6 and Bartlett’s test was significant (p < 0.001), the dataset was considered adequate to perform rPCA (Tabachnick & Fidell, 2013; Kaiser & Rice, 1974). (2) *Component extraction and estimation*. Principal component method was used to extract a smaller number of components that capture the most variance of the initial variables, representing the common cognitive processes underlying the 13 original cognitive measures. Scree plots and parallel analysis were used to determine the number of components (Horn, 1965). Parallel analysis generated a set of random correlation matrices (n = 500) by using the same number of variables (n = 13) and participants (N = 461) in the parallel analysis engine (https://analytics.gonzaga.edu/parallelengine/). Then, we compared the solution to the random simulated normal data. The eigenvalues obtained from the actual matrix were compared with those obtained from the randomly generated matrices, and the estimated number of components was determined by selecting components with eigenvalues larger than 95^th^ percentile of the randomly generated components’ eigenvalues (Figure 2a). (3) *Rotation*. To examine the component structure among the initial variables, an orthogonal rotation method Varimax that constrains the components to be uncorrelated was applied to the coefficient matrix (Kaiser, 1958). (4) *Component coefficients*. Component coefficients (*r*), reflecting the correlation between each of the components and each of the original variables, were thus obtained. Given the sample size in this study (N > 200), component coefficients that were larger than 0.4 were deemed practically significant (Hair Jr et al., 1998; Stevens, 1992). (5) *Component scores*. Component scores were calculated by a regression method where the regression weights were found from the inverse of the correlation matrix times the coefficient of each variable on the corresponding components. The coefficients were used to interpret the components, and component scores were used for the following analyses. In subsequent analyses, we used these cognitive components, rather than individual tests to measure the impacts of risk factors and lifestyle activities on cognition.

**Figure 2.**
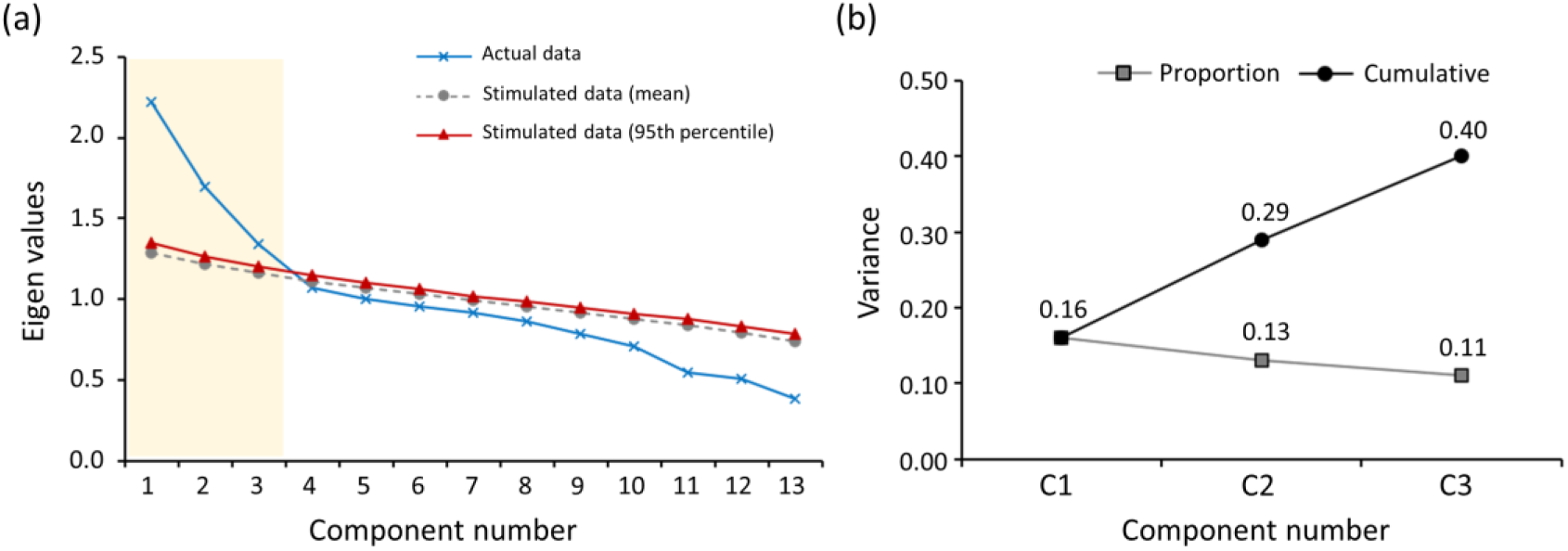
Data reduction of cognitive information. (a) Scree plot and parallel analysis. Eigenvalues of the principal components obtained from the actual data (in blue line) were compared to those of random simulated data (mean values in grey line and 95^th^ percentile in red line). Eigenvalues larger than 95^th^ percentile of the randomly generated 500 eigenvalues (simulation) determined the number of components. (b) The three extracted components were rotated to be uncorrelated with each other. The proportion of variance explained by each component, and the cumulative variance explained by the three components are presented for the cognitive analysis cohort. Abbreviations: C1 = component 1; C2 = component 2; C3 = component 3.

### Statistical approach

We used the SPSS and R software for all statistical analyses. The normality of the data was assessed by combining the visualization of a quantile-quantile plot and the Shapiro–Wilk test. Demographic and clinical information of the study cohort was analyzed across risk groups using chi-square (χ^2^ tests) for categorical variables and Mann-Whitney U tests for continuous (discrete) variables, given that they were not normally distributed in this cohort.

To investigate the effects of lifestyle activities and risk factors on cognition and moderating role of the sex variable, we used hierarchical regression models to look at the contribution of mid-life lifestyle factors (LEQ specific and non-specific scores), risk factors (*APOE* _Ɛ_4 genotype and FHD) and their interactions with sex on cognitive performance. Additionally, to investigate how risk factors impact the associations of sex by lifestyle interaction term and cognition, the participants were stratified into low- or high-risk groups based on APOE genotype, FHD and CAIDE risk score. CAIDE score was treated as a binary variable, where a median split (CAIDE score = 6) was used to divide participants into low or high CAIDE groups. For each group, we used multiple regression models to look at the contribution of lifestyle activities (LEQ specific and non-specific scores) and their interactions with sex on cognitive performance separately.

The effect of APOE _Ɛ_4 and CAIDE risk factors was modelled independently, in order to avoid modelling the variance associated with APOE genotype in the same model twice. In each case, dependent variables were the aforementioned composite cognitive components, each assessed in a separate model.

To avoid multicollinearity, we mean-centered continuous variables (specific and non-specific scores). Age, sex and years of education were included as covariates in hierarchical regression models. A significant interaction effect between a risk/ lifestyle factor and sex would indicate that the effect of risk/lifestyle factor on cognitive performance differs across males and females. For any observed interactions, we plotted the regression of the risk/lifestyle factor on cognitive performance for males and females, to interpret the effect. We then tested the significance of the slopes of the simple regression lines, to investigate for males or females, whether we found an effect of risk/lifestyle factor on cognitive performance. Scatter plots showing the relationship between cognitive performance and lifestyle factors were generated using unadjusted values, and full statistical details are provided for reference in each legend.

## Results

### Demographic characteristics

491 participants were recruited from four study sites. All 491 completed cognitive and clinical assessments. Mild cognitive impairment and dementia were ruled out based on detailed clinical assessment at each site. Demographic specifications of the cohort, stratified by APOE _Ɛ_4 genotype and family history of dementia, are shown in Table 1. There were no significant differences in age, sex or years of education between the groups. As expected, APOE _Ɛ_4 allele genotype was more frequently found in the FHD+ than FHD- group (p = 0.01). Naturally, CAIDE scores including APOE status were significantly higher for the APOE _Ɛ_4+ than APOE _Ɛ_4-group (p < 0.001), but when APOE status was excluded the CAIDE scores did not differ between the two groups (Table 1).

### Cognitive domains

The challenge of using cognitive tests that capture subtle changes in information processing in midlife due to dementia risk, that may not conform to domains of impairment in clinical populations, was addressed here with a novel multidimensional cognitive assessment battery. We used a data-driven approach to reduce the cognitive data into the most relevant aspects that could maximize power to detect subtle cognitive changes. We first performed parallel analysis and plotted scree plot to determine the number of components that best represented the original 13 cognitive measures. Results showed that eigenvalues of the first three components were larger than 95^th^ percentile of the randomly generated eigenvalues (Figure 2a), which indicated that the three component (C) solution best represented the data. The three components were then rotated to be uncorrelated with each other, and could cumulatively explain a total of 40% percentage of the variance (C1 = 16%, C2 = 13%, C3 = 11%) (Figure 2b).

The loading values (weights) of the cognitive measures reflect the relationships between the original measures and the corresponding components, shown in Figure 3 for the Imperial College London dataset (Figure 3a [from Deng et al., 2022]) and the other four sites (Figure 3b), for comparison. Measures with higher coefficients were more closely related to the components. The components can be interpreted by mapping out the cognitive functions that the highest loading measures tapped into (Figure 3; Supplementary Table 2). The four measures that load strongly on C1 reflect memory recall with a strong episodic element (3/4: descriptive recall, narrative recall, name-face association [recall of relations]) in different modalities (i.e., verbal, visuo-spatial). For subsequent analyses, we refer to C1 as ‘episodic and relational memory.’ The four measures that load strongly on C2 reflect processing of visual, auditory/linguistic and multisensory integration of audio-visual information. For subsequent analyses, we refer to C2 as ‘multisensory processing.’ The two measures that load strongly on C3 reflect the VSTMB task, which involved recalling of single features (shape) or feature binding (shape and colour). For subsequent analyses, we refer to C3 as ‘short-term memory binding.’ For full description of each task, please see Supplementary Table 2.

**Figure 3.**
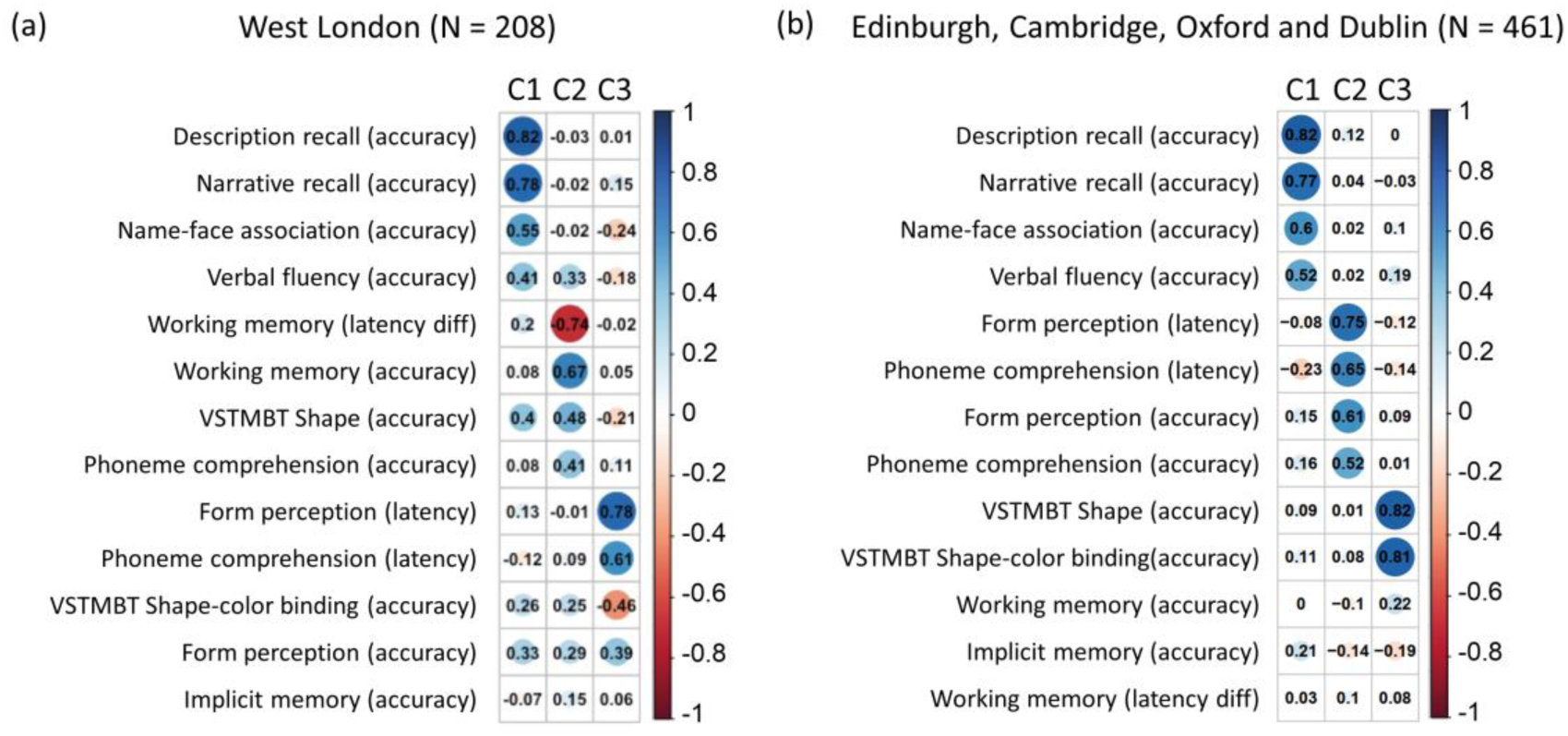
Component loadings from (a) the West London site, and (b) the Edinburgh, Cambridge, Oxford and Dublin sites. The loading of each cognitive measure (in rows) on each cognitive component (in columns) are shown. A larger absolute coefficient (darker colors and larger solid circles) represents a closer relationship between the cognitive measure and the corresponding component. Cool/warm colors represent the positive/negative relationships between cognitive measures and components, as shown in the color-bar scale. The color-bar indicates loading values, higher loading values indicate stronger contribution to corresponding components. Abbreviations: C, component; VSTMBT, visual short-term memory binding test; diff, difference. The component loading from the West London site was adapted from Deng et al., 2022.

Comparison to Deng et al. (2022). The measures that loaded on the first component replicated those in the previous study. The top measures that loaded on the other two did not replicate findings from the previous study. By contrast to Deng et al. (2022), within each of the bottom two component, the top measures cohered with one another – they represented the same cognitive function. These findings validated the data-reduction method, in that it carved up the cognitive data into three coherent and distinct cognitive spaces – episodic and relational memory, multisensory processing, and short-term memory binding – two of which (episodic and relational memory (Bäckman et al., 2001) and short-term memory binding [Parra et al., 2016; Parra et al., 2017]) are known to be very vulnerable to early AD processes in mild cognitive impairment. The first component being the same as in Deng et al. (2022), provides very strong evidence that episodic and relational memory is the top cognitive function undergoing performance variability in mid-life. The differing and stronger results here relative to Deng et al. (2022) for the other two components are likely due to the larger sample (N = 461 vs N = 208) yielding higher power to detect the underlying distribution of pattern performance variability in this age group.

### Effect of risk factors on cognition

#### Episodic and relational memory

##### APOE

The multiple linear regression model showed no associations between APOE _Ɛ_4 and episodic and relational memory. There was a significant effect of sex, with females performing better than males [β (SE) = 0.51 (0.09), p < 0.001] (Table 2, Figure 6a). This effect was independent of age and years of education. Higher education values were significantly associated with better performance in episodic and relational memory [β (SE) = 0.04 (0.01), p = 0.003]. The inclusion of APOE _Ɛ_4 by sex interaction terms (APOE _Ɛ_4 × sex) into the hierarchical regression model (Supplementary Table 3a) showed a trend association between APOE _Ɛ_4 x sex interaction term and episodic and relational memory [β (SE) = 0.35 (0.18), p = 0.06].

**Table 2.** APOE – Regression coefficients for Episodic and Relational Memory (Component 1)

| Model summary |  | R<sup>2</sup> | F | p |
| --- | --- | --- | --- | --- |
|  |  | 0.10 | 12.89 | <0.0001 |
| DV | IV | β (SE) | p |  |
| Component 1 | APOE ε4 | 0.07 (0.09) | 0.44 |  |
|  | Age | -0.02 (0.01) | 0.07 |  |
|  | Sex | 0.51 (0.09) | <0.001 |  |
|  | Years of education | 0.04 (0.01) | 0.003 |  |

Note: unstandardized coefficients β and standard error (SE) were reported. DV, dependent variable; IV, independent variable; APOE ε4, Apolipoprotein ε4.

##### CAIDE

The hierarchical regression model showed a significant negative association between CAIDE score and episodic and relational memory [β (SE) = -0.05 (0.02), p = 0.003] (Table 3, Figure 4). Higher CAIDE scores were significantly associated with worse performance. This result was not driven by APOE status, since the relationship stood when CAIDE score without APOE status was included as the independent variable (Supplementary Table 3b). There was a trend association between CAIDE score and episodic and relational memory on 208 participants from the West London site [β (SE) = -0.05 (0.03), p = 0.057] (Supplementary Table 3c). No interaction between CAIDE and sex on episodic and relational memory was observed (Supplementary Table 3d).

**Table 3.** CAIDE – Regression coefficients for Episodic and Relational Memory (Component 1)

| Model summary | | $R^2$ | F | p |
| --- | --- | --- | --- | --- |
|  |  | 0.02 | 9.20 | 0.003 |
| DV | IV | $\beta$ (SE) | p | |
| Component 1 | CAIDE | -0.05 (0.02) | 0.003 |  |

Note: unstandardized coefficients $\beta$ and standard error (SE) were reported. DV, dependent variable; IV, independent variable; CAIDE, Cardiovascular Risk Factors, Aging and Dementia.

**Figure 4.**
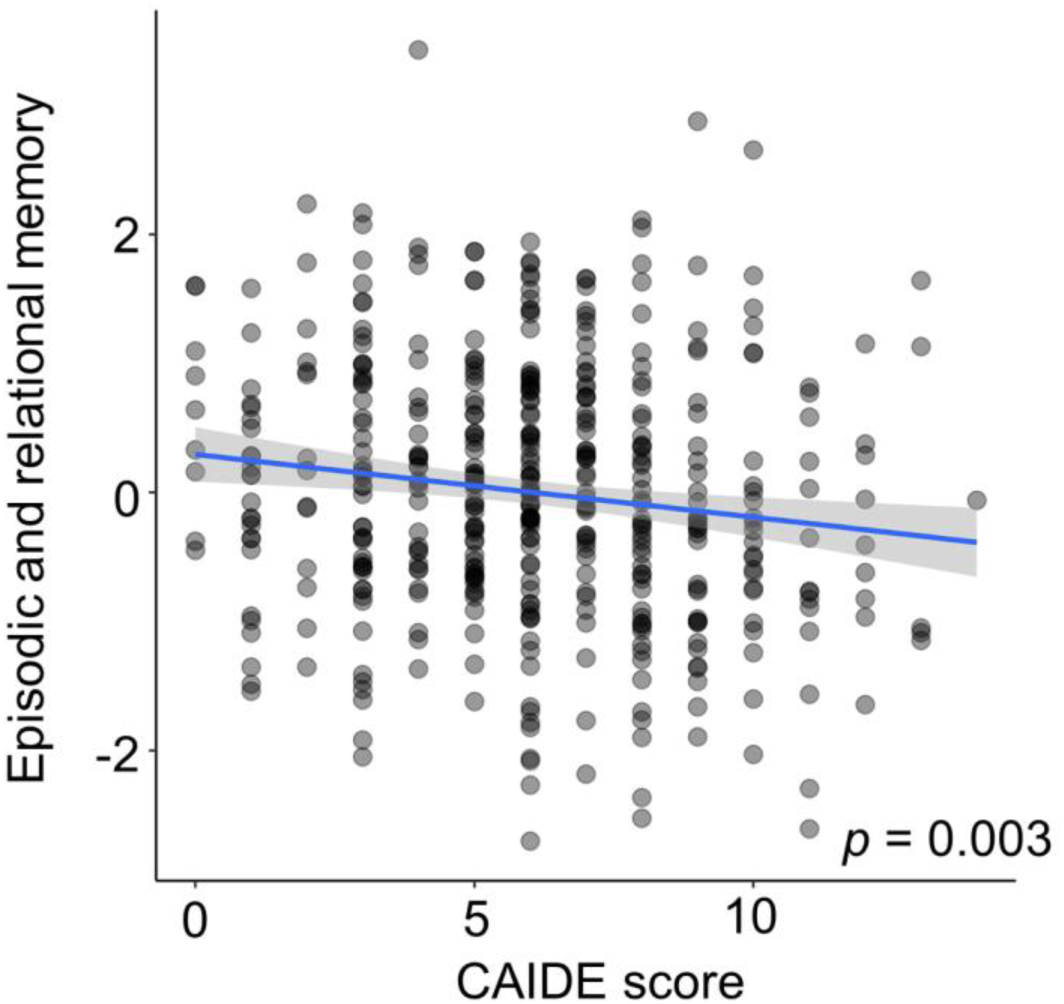
Association of the CAIDE score with episodic and relational memory performance. On the x axis, higher scores represent higher CAIDE score, and on the y axis, higher scores represent better episodic and relational memory performance. Scatter plots show unadjusted values. Full regression statistic for CAIDE: [β (SE) = - 0.05 (0.02), p = 0.003]. Abbreviation: CAIDE, Cardiovascular risk factor, Aging, and Incidence of Dementia.

##### FHD

There were no significant associations between FHD with episodic and relational memory (Supplementary Table 3e).

The other two cognitive components (multisensory processing and short-term memory binding) showed no significant relations to risk factors (Supplementary Table 4a, 4b, 4c; Supplementary Table 5a, 5b, 5c).

### Effect of lifestyle factors on cognition

Comparison to Heneghan et al. (2022). We then investigated the effect of lifestyle factors on episodic and relational memory. The hierarchical regression model showed a significant positive association between the non-specific LEQ factor and episodic and relational memory [β (SE) = 0.06 (0.01), p < 0.001] (Table 4, Figure 5). More frequent engagement in physically, socially and intellectually stimulating activities was associated with better episodic and relational memory, independent of age, sex, and years of education. This effect replicated the result reported by Heneghan et al. (2022).

**Table 4.** Lifestyle – Regression coefficients for Episodic and Relational Memory (Component 1)

| Model summary | | $R^2$ | F | p |
| --- | --- | --- | --- | --- |
|  |  | 0.14 | 14.74 | <0.001 |
| DV | IV | $\beta$ (SE) | p | |
| Component 1 | Specific | 0.01 (0.01) | 0.29 |  |
|  | Non-specific | 0.06 (0.01) | <0.001 |  |
|  | Age | -0.02 (0.01) | 0.02 |  |
|  | Sex | 0.51 (0.09) | <0.001 |  |
|  | Years of education | 0.03 (0.01) | 0.046 |  |

Note: unstandardized coefficients $\beta$ and standard error (SE) were reported. DV, dependent variable; IV, independent variable.

**Figure 5.**
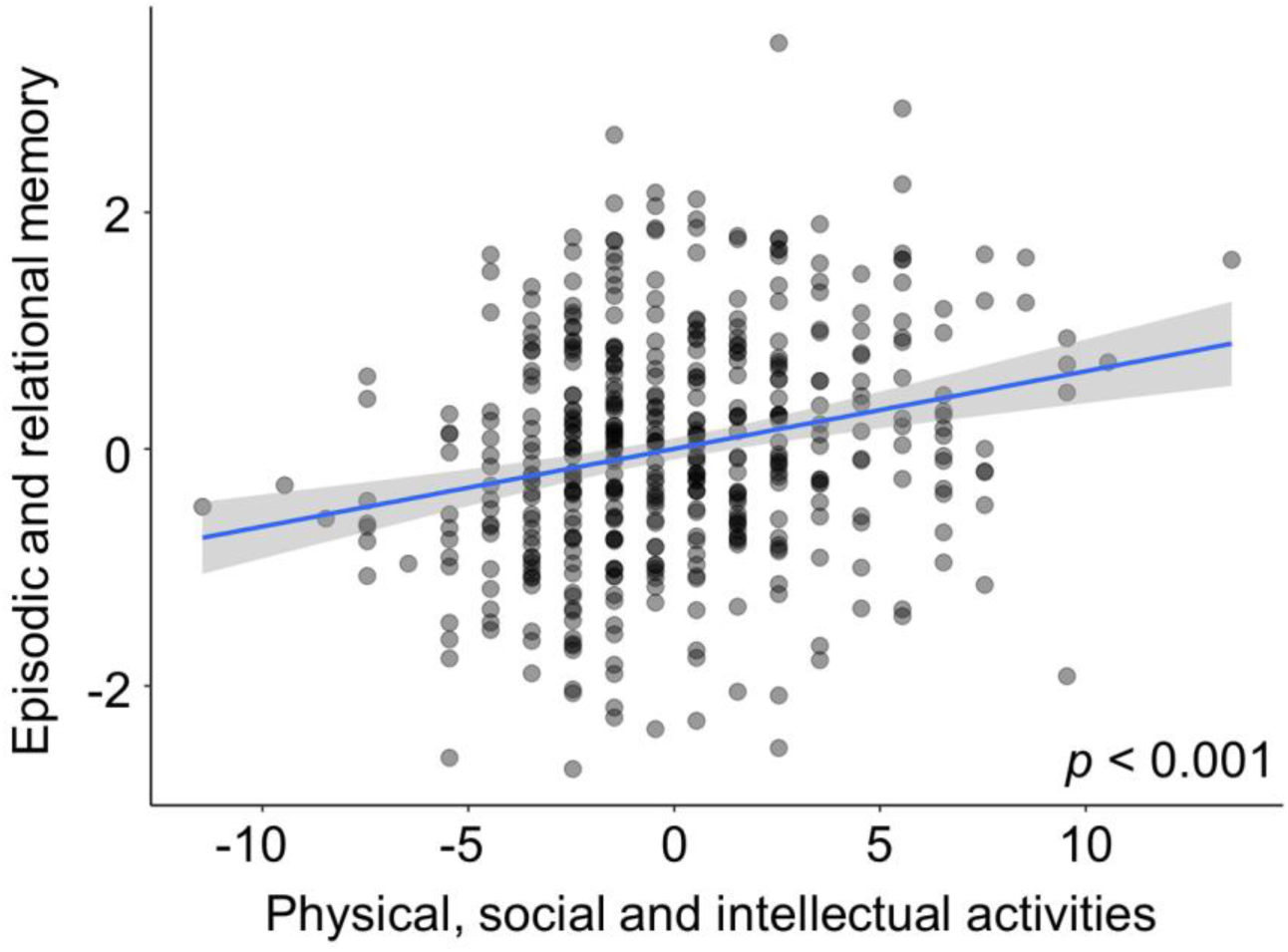
Association of physical, social and intellectual activities with episodic and relational memory performance. On the x axis, higher scores represent more frequent engagement in physical, social and intellectual lifestyle activities, and on the y axis, higher scores represent better episodic and relational memory performance. Scatter plots show unadjusted values. Full regression statistic for physical, social and intellectual activities, after controlling for sex, age and education: [β (SE) = 0.06 (0.01), p < 0.001]

Interactions of risk factors, stimulating lifestyle activities and biological sex Given the strong effect of sex on episodic and relational memory (Figure 6a), we looked at potential interactions between sex, risk factors and lifestyle factors on cognition. The inclusion of sex by lifestyle interaction terms (sex × specific score, sex × non-specific score) into the hierarchical regression model revealed a significant association between the specific LEQ score x sex interaction term and cognitive performance [β (SE) = 0.04 (0.02), p = 0.035] (Table 5). To interpret this interaction, we investigated the relationship between specific lifestyle activities and cognition for females and males independently. We found a significant relationship between specific lifestyle activities and cognition in females [β (SE) = 0.03 (0.01), p = 0.03], but not males [β (SE) = -0.01 (0.02), p = 0.43] (Figure 6b). These results suggested that females with higher occupational attainment performed better in episodic and relational memory.

**Figure 6.**
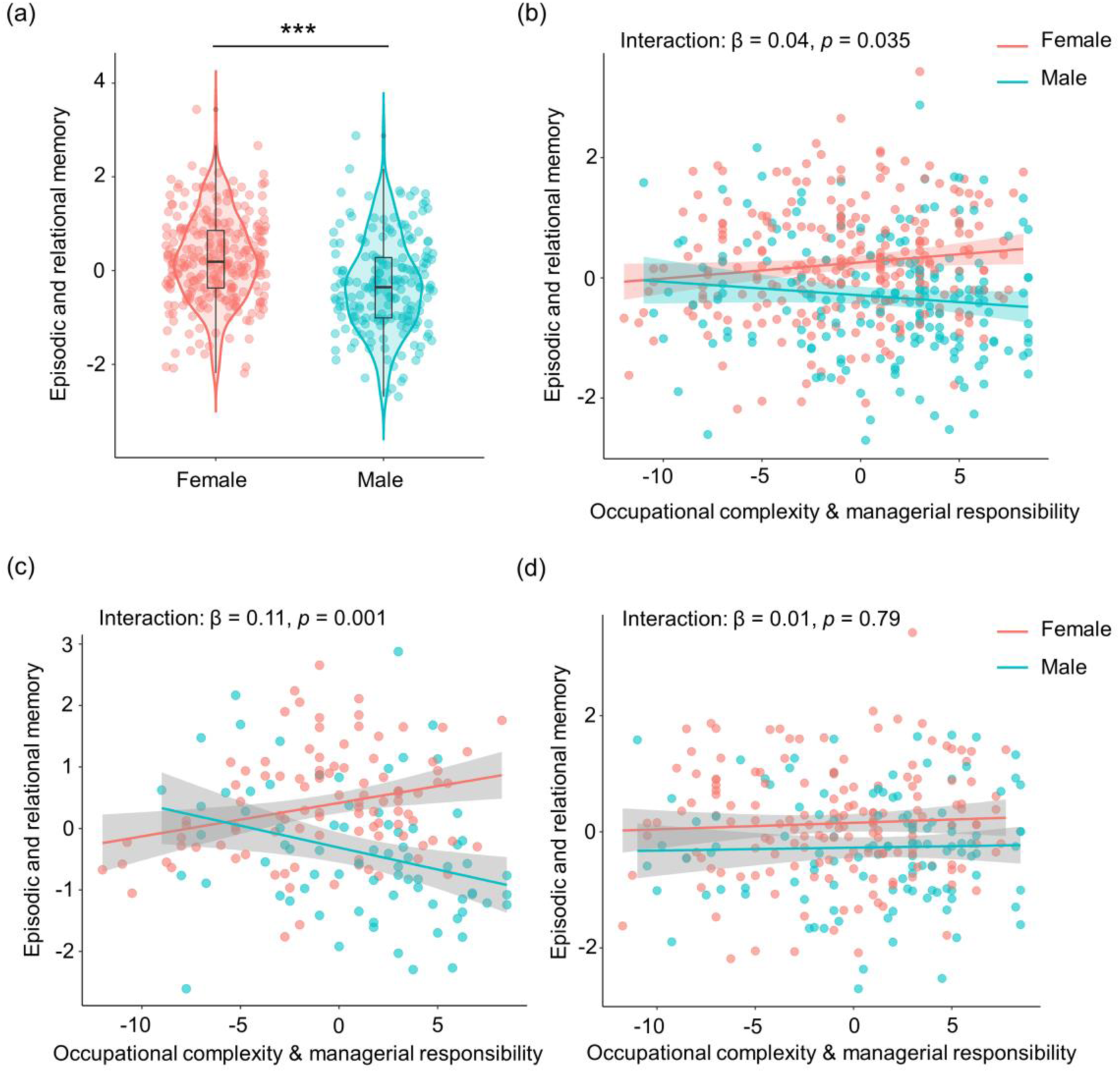
Sex differences in cognition and in the associations between risk, protective lifestyle factors and cognition in mid-life. On the x axis, higher scores represent greater occupational complexity and managerial experience, and on the y axis, higher scores represent better episodic and relational memory performance. (a) Sex difference on episodic and relational memory. Females performed better than males in episodic and relational memory. \*\*\**p* < 0.0005. Interactions of occupational complexity and managerial responsibility and sex on episodic and relational memory in (b) the four sites cohort; (c) in the APOE _Ɛ_4+ group; and (d) in the APOE _Ɛ_4-group. In the four sites cohort, females showed a significant positive association between occupational complexity and managerial responsibility and episodic and relational memory. No significant association was observed for males. In APOE _Ɛ_4+ group, males showed a significant negative association between occupational complexity and managerial experience and episodic and relational. A trend positive association was seen for females. In APOE _Ɛ_4-group, there was no significant interaction of occupational complexity and managerial responsibility and sex on episodic and relational memory. Occupational complexity and managerial responsibility are mean-centred. The scatter plot shows unadjusted values, but the statistical significance was based on the regression analyses where we controlled for covariates.

**Table 5.**
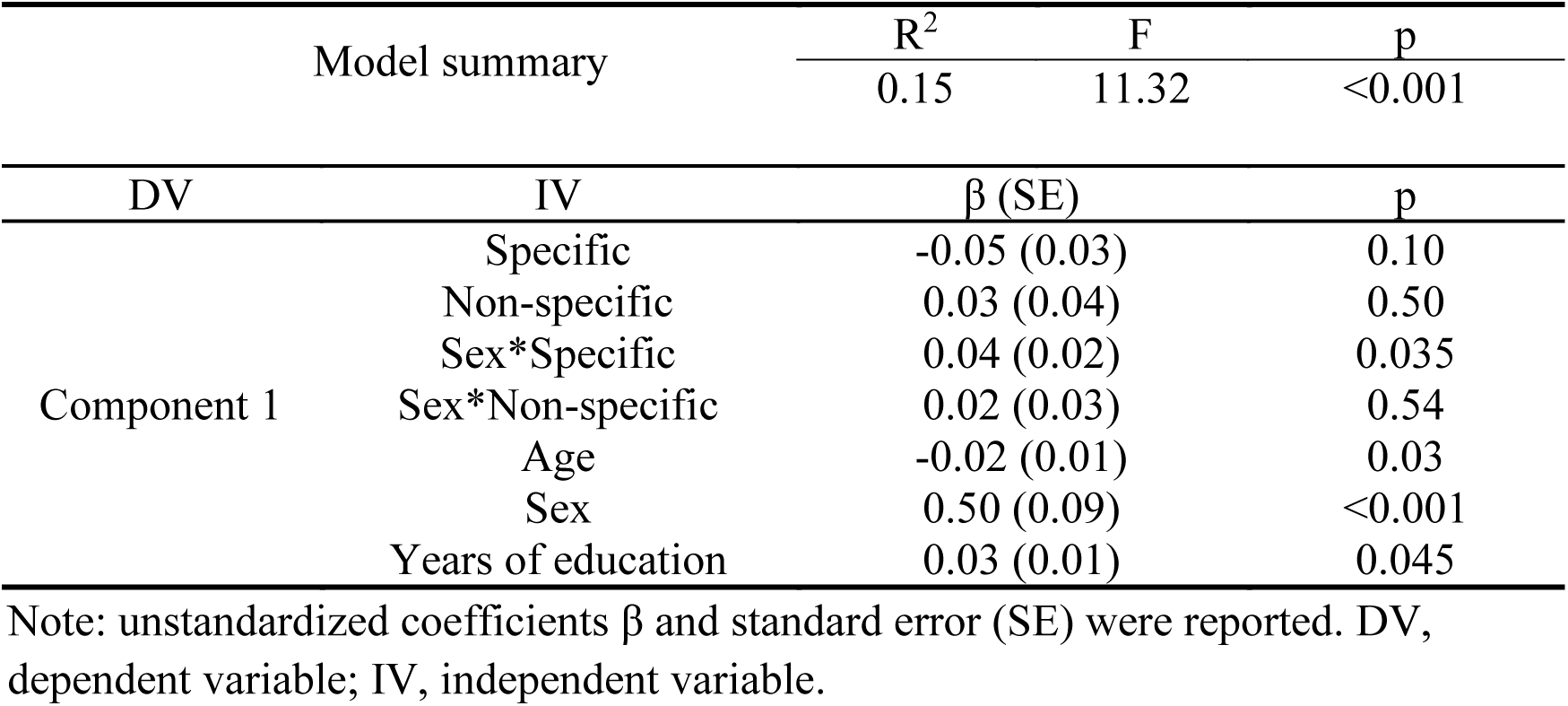
Lifestyle & Sex – Regression coefficients for Episodic and Relational Memory (Component 1) including interaction terms

| Model summary |  | R<sup>2</sup> | F | p |
| --- | --- | --- | --- | --- |
|  |  | 0.15 | 11.32 | <0.001 |
| DV | IV | $\beta$ (SE) | | p |
| Component 1 | Specific | -0.05 (0.03) |  | 0.10 |
|  | Non-specific | 0.03 (0.04) |  | 0.50 |
|  | Sex*Specific | 0.04 (0.02) |  | 0.035 |
|  | Sex*Non-specific | 0.02 (0.03) |  | 0.54 |
|  | Age | -0.02 (0.01) |  | 0.03 |
|  | Sex | 0.50 (0.09) |  | <0.001 |
|  | Years of education | 0.03 (0.01) |  | 0.045 |

Note: unstandardized coefficients $\beta$ and standard error (SE) were reported. DV, dependent variable; IV, independent variable.

Additionally, we examined how risk factors impacted these interactions. There were significant associations between sex by lifestyle interaction terms (sex × specific score, sex × non-specific score) and episodic and relational memory in groups stratified by APOE genotype. For the APOE _Ɛ_4+ group, we observed a significant positive association between the specific LEQ score x sex interaction term and episodic and relational memory [β (SE) = 0.11 (0.03), p = 0.001] (Table 6). To interpret this interaction, we investigated the relationship between specific lifestyle activities and cognition for females and males independently (Figure 6c). We found a significant relationship between specific lifestyle activities and cognition in males [β (SE) = -0.06 (0.03), p = 0.01], and a trend association between specific lifestyle activities and cognition in females [β (SE) = 0.04 (0.02), p = 0.06]. For APOE _Ɛ_4 carriers, greater occupational complexity and managerial responsibility was associated with worse cognition in males, and stronger (trend effect) cognition in women. For the APOE _Ɛ_4-group, we did not observe a significant association between lifestyle x sex interaction term and episodic and relational memory (Supplementary Table 6a, Figure 6d). Additionally, there were no significant associations between interactions of sex x lifestyle and cognition in FHD+/FHD-groups (Supplementary Table 6b, 6c) or high CAIDE/low CAIDE groups (Supplementary Table 6d, 6e).

**Table 6.** Lifestyle & Sex – Regression coefficients for Episodic and Relational Memory (Component 1) including interaction terms in APOE _Ɛ_4+ group

| Model summary | | $R^2$ | F | p |
| --- | --- | --- | --- | --- |
|  |  | 0.25 | 8.15 | <0.001 |
| DV | IV | $\beta$ (SE) | | p |
| Component 1 | Specific | -0.17 (0.05) |  | 0.002 |
|  | Non-specific | 0.04 (0.07) |  | 0.56 |
|  | Sex*Specific | 0.11 (0.03) |  | 0.001 |
|  | Sex*Non-specific | 0.001 (0.04) |  | 0.85 |
|  | Age | -0.01 (0.01) |  | 0.69 |
|  | Sex | 0.71 (0.14) |  | <0.001 |
|  | Years of education | 0.02 (0.02) |  | 0.42 |

Note: unstandardized coefficients $\beta$ and standard error (SE) were reported. DV, dependent variable; IV, independent variable; APOE $\epsilon 4+$ , Apolipoprotein $\epsilon 4$ genotype positive.

## Discussion

It is now acknowledged that Alzheimer’s Disease processes are present decades before the onset of clinical symptoms (Ritchie et al., 2015; Jack Jr et al., 2010), but, until recently, it had remained unknown whether lifestyle factors could protect against the impact of risk factors in mid-life, prior to the development of dementia. Heneghan et al. (2022) showed that modifiable lifestyle factors start to offset the impact of AD risk (i.e., family history of dementia) on cognition from mid-life (N = 208). A trend effect of sex was found on episodic and relational memory, but the effect of sex on the relationship between risk, lifestyle factors and cognition was not investigated. Sex is an important biological variable in the development of AD (Udeh-Momoh and Watermeyer, 2021; Luckey et al., 2021). Strikingly, however, very little is known on its moderating effect on these relationships in mid-life. Our objectives in this study were twofold. First, we aimed to replicate our recent findings [Deng et al. (2022) and Heneghan et al. (2022)] on the contribution of mid-life modifiable activities to cognition in individuals with risk of late-life dementia, in a larger cohort (N = 461) of the PREVENT–Dementia research program. Second, we investigated associations between biological sex, dementia risk, protective lifestyle factors and cognitive performance.

We found that the top dimension of cognitive variability in this cognitively unimpaired cohort comprised episodic and relational memory performance, replicating our previous finding (Deng et al., 2022). Furthermore, we found that performance in this domain was both negatively associated with the CAIDE dementia risk and positively associated with engagement in physically, socially and intellectually stimulating activities in mid-life. This effect was independent of sex, age, years of education and cardiovascular factors captured by the CAIDE score. Furthermore, we also replicated a trend effect of sex on this cognitive domain, showing that females performed significantly better than males in episodic and relational memory (Heneghan et al., 2022). This result is consistent with previous studies showing that women typically outperform healthy men on episodic memory tests (Asperholm et al., 2019). The replication of our previous results (Deng et al., 2022; Heneghan et al., 2022) provided very strong evidence that episodic and relational memory are potentially the earliest (from mid-life) cognitive functions to be impacted by dementia risk in cognitively unimpaired individuals.

The key novel question in this study, however, was to investigate whether sex modulated the relationship between dementia risk, protective lifestyle activities and cognition. The first novel finding of this study was that sex modulated the relationship between occupational attainment and cognition. Only for females, not males, was higher occupational attainment in mid-life associated with better episodic and relational memory. Importantly, this effect was independent of age, years of education, and other non-occupation related lifestyle activities.

The second novel finding was that inherited dementia risk (i.e., APOE _Ɛ_4 carriership) modulated the association between sex, protective lifestyle activities and cognition. For female APOE _Ɛ_4 carriers, higher occupational attainment in mid-life was associated with better (trend level) episodic and relational memory. Conversely, for male APOE _Ɛ_4 carriers, occupational attainment in mid-life was associated with worse episodic and relational memory. These interaction effects of sex by occupational attainment were absent for female/male APOE _Ɛ_4 non-carriers. This effect was not driven by an interaction of APOE _Ɛ_4 x sex on cognition alone, because APOE _Ɛ_4 carriership was not associated with differential cognition in males, and only a weak effect of APOE on cognition was seen in females.

To the best of our knowledge, this is the first study showing a robust interaction between biological sex, genetic risk (APOE _Ɛ_4 carriership), and lifestyle factors (occupational attainment) on cognition in middle-aged cognitively unimpaired individuals. A key result from this interaction is that higher occupational attainment in female APOE _Ɛ_4 carriers was associated with stronger cognition.

Why do female but not male APOE _Ɛ_4 carriers appear to benefit cognitively from occupational attainment in mid-life? Previous studies show a sex-specific effect of APOE _Ɛ_4 allele, with increased risk of tau-related neurodegeneration in female relative to males APOE _Ɛ_4 carriers (Mofrad et al., 2020; Hohman et al., 2018). It has been suggested that APOE _Ɛ_4 serves to exacerbate small discrepancies between males and females in disease pathogenesis, such as through tau deposition and related neurodegeneration, that compound in the later stages of disease (Udeh-Momoh and Watermeyer, 2021). In addition to greater vulnerability to AD through the impact of sex-specific APOE _Ɛ_4 carriership and multi-system vulnerabilities set in motion by the menopausal transition in mid-life (Udeh-Momoh and Watermeyer, 2021), females traditionally have accrued higher vulnerability to neurodegeneration relative to males through barriers to key reserve contributors, such as educational and occupational opportunities (Russ et al., 2013). These multidimensional vulnerability influences underlie recent findings that women benefit from reserve contributors more than men (Subramaniapillai et al., 2021).

What might be the mechanism by which occupational attainment impacts cognition in mid-life? Models of AD disease delineated by neuropathological staging (Braak & Braak, 1991) place the locus coeruleus, a small nucleus in the pontine tegmentum region of the brainstem, at the pathogenesis of AD (Jacobs et al., 2021; Arnsten et al., 2021; Márquez & Yassa, 2019). The LC is responsible for the production of neurotransmitter noradrenaline (NA) (Amaral & Sinnamon, 1997), a major driver of the brain’s arousal system, which strongly modulates high-order cognition. Novelty and complexity, key features associated with high-skill occupations involving leadership roles, provide triggers for arousal, and, thus, the LC–NA activity occurs strongly during engagement in these roles. Therefore, such job activities upregulate the LC and may protect it from early neurodegeneration due to pTau deposition. Furthermore, as the brain site of noradrenaline, the LC is key to the protective effect of lifestyle factors on the whole brain. Studies suggest that environmental enrichment, such as the mentally stimulating occupational roles measured in this study, upregulates the noradrenergic system, which otherwise depletes with age (Liu et al., 2020) and AD pathology (Jacobs et al., 2021), leading to compensatory brain mechanisms for cognitive function, such as the strengthening of the fronto-parietal brain and other large-scale brain networks (Robertson, 2014; Robertson, 2013).

In summary, our results suggest that modifiable lifestyle activities, particularly occupational attainment, may offset cognitive decrements in APOE _Ɛ_4 carriers females. Occupational attainment is a key reserve contributor (Stern et al., 2020; Stern, 2012) and has been positively associated with late-life cognition and decreased dementia symptoms (Bosma et al., 2003; Andel et al., 2015; Smart et al., 2014; Huang et al., 2020). Our finding advances understanding by showing that occupational attainment specific to mid-life contributes to cognitive reserve against inherited risk of dementia, or even incipient AD neuropathology, in females who are presently cognitively healthy but carry the APOE _Ɛ_4 allele. These findings point to an urgent need for understanding the underlying mechanisms associated with divergent effects of lifestyle factors in mid-life, in females versus males with inherited risk of dementia. Finally, as women disproportionally to other life stages leave work in mid-life (New York Times, 2014) due to increased caregiving duties –childrearing and caretaking of ageing parents – our results suggest that retention of women in workplace is important for protecting brain health in a high-risk population, and, thus, needs to supported with appropriate policy development.

### Methodological considerations

As education is strongly positively linked to IQ (Ritchie & Tucker-Drob, 2018) and our cohort was highly educated, the question of reverse causation arises (Chan et al., 2018; Gow et al., 2017; Borgeest et al., 2020). This points to the possibility that better education and resulting high cognitive activities may determine high occupational attainment, rather than the inverse. However, the effect on mid-life episodic and relational memory was independent of the total years of education, which shows that education does not directly drive this effect. Furthermore, we found that mid-life occupational attainment was associated with improved cognitive performance only in APOE _Ɛ_4 carrier females, which were not, at least not prima facie, more educated than APOE _Ɛ_4 non-carrier females. Conversely, APOE _Ɛ_4 carrier men did showed a negative association of occupational attainment with cognition. Therefore, this interaction effect is likely independent of any indirect effects of education. Another instantiation of reverse causation could that APOE _Ɛ_4 carriership may cause some benefit to cognition of females, leading them to end up in more highly skilled occupations and to progress in managerial responsibilities. There was no evidence for a major benefit of APOE _Ɛ_4 carriership on cognition in females in our data.

## Supporting information

SupplementaryInformation_Qi_etal_2023

## Data Availability

All data produced in the present study are available upon reasonable request to the authors

## Acknowledgement

Q.Q. was funded by the China Scholarship Council—Trinity College Dublin Joint Scholarship Programme. L.N. was funded by a L’Oréal-UNESCO for Women In Science International Rising Talent Award, the Welcome Trust Institutional Strategic Support grant, and the Global Brain Health Institute Project Grant.

This work was funded by grants for the PREVENT-Dementia program from the UK Alzheimer’s Society (Grant nos. 178, 264 and 397). The PREVENT-Dementia study is also supported by the US Alzheimer’s Association (Grant no. TriBEKa-17–519007) and philanthropic donations.

We thank all PREVENT-Dementia participants for their enthusiastic participation in this study, the DeNDRoN specialty within the Clinical Research Network.

