## SupplementaryInformation_Qi_etal_2023 for "Sex differences in the associations between risk for late-life AD, protective lifestyle factors and cognition in mid-life"

**Title**

**Materials and Methods**

The eleven cognitive summary variables from the COGNITO battery were:

1. Working memory: the simultaneous presentation of auditory and visual attention tasks assessed by subtracting the time taken in milliseconds on this double task from the visual task alone.
2. Working memory: the simultaneous presentation of auditory and visual attention tasks assessed by total number of correct answers for visual form recognition on this double task.
3. Narrative recall: total number of correct elements on immediate recall of a story with a temporal progression requiring attention to macrostructure.
4. Description recall: total number of correct elements recalled of a description without thematic progression requiring attention to microstructure and recall of spatial location. The narrative and description recall are similar in terms of word frequency in the language and syntactic structure.
5. Implicit memory: difference in the number of steps in the progressive build-up of names on the screen required for recognition between names never seen and number of names previously learnt in an immediate recall task.
6. Name-face association: number of faces recognized after a delay from a series of 18 faces of which 9 have been previously shown with their corresponding names.
7. Form perception: number of correct answers in the matching of complex forms to a multiple-choice array.
8. Form perception speed: mean time taken in milliseconds for each trial.
9. Phoneme comprehension: number of correct responses in the matching of a word with an image presented as part of a multiple-choice array including semantic, morphological and phonetic distractors.
10. Phoneme comprehension speed: mean time taken in milliseconds to perform.
11. Verbal fluency: total sum of the number of words generated in 30s using both a semantic (vegetables) and phonemic (letter P) cue.

The two summary variables from the Visual Short-term Memory Binding task were:

12. Visual short-term memory binding test (shape only condition): percentage of correct recognition of the shape of presented visual stimuli after a short period of retention.
13. Visual short-term memory binding test (shape-color binding condition): percentage of correct recognition of combinations of shape and color of presented visual stimuli after a short period of retention.

**Behavioral data reduction**

Sharp breaks in the “scree" plot of the successive eigenvalues suggest the appropriate number of components to extract. We also conducted a parallel analysis that compares the scree of components of the actual data with that of a random data matrix of the same size as the original (Horn, 1965). The detailed steps are: 1) We simulated a random normal data matrix of the same number of original cognitive variables (n = 13) and the same number of participants (N = 461). 2) We extracted eigenvalues from the simulated data matrix. We repeated these two steps 500 times to create a set of 500 parallel eigenvalues. 3) We took the mean and 95th percentile of all eigenvalues generated by principal components analysis of random data sets. The results were a vector of mean (and 95th percentile) eigenvalues equal in size to the original number of cognitive variables (n = 13). 4) We compared the eigenvalues of the actual data to that of parallel random data sets. Specifically, we plotted eigenvalues from the actual and random data sets and kept only those components whose eigenvalues are greater than 95th percentile of eigenvalues from the random data sets. Figure 2a in the manuscript shows the scree plots of eigenvalues based on the actual data matrix (in blue line) and simulated data matrices. The mean eigenvalues are in dashed grey line, and the 95th percentile of eigenvalues are in red line. The shaded areas indicate that the eigenvalues of components based on the actual data were larger than the mean and 95th percentile of eigenvalues from the random data sets. Figure 2b shows the proportion of variance that each component explained, and the cumulative variance of the three components.

**Supplementary Figures**

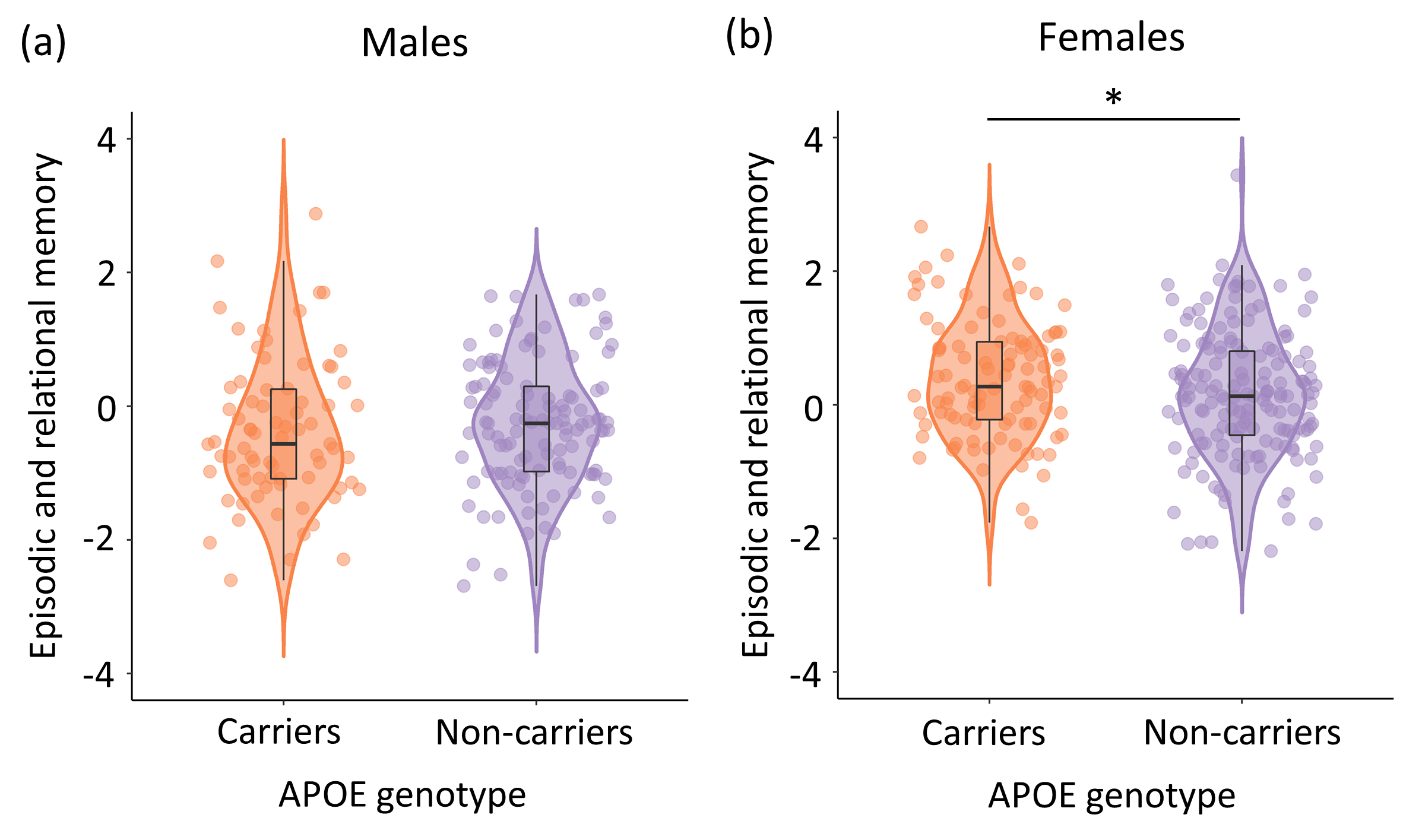
Supplementary Figure 1. The effect of APOE genotype on episodic and relational memory in (a) males and at (b) females. Full regression statistic for APOE genotype, after controlling for age and education: [Males: β (SE) = -0.15 (0.15), p = 0.31; Females: β (SE) = 0.23 (0.12), p = 0.045]. **p* < 0.05.

**Supplementary Tables**

Supplementary Table 1. Complete list of risk factors and tests obtained in the cohort

| PREVENT–Dementia program four sites | |
| --- | --- |
| **Risk factors** (Missing = 10) | |
| *Family history* | |
| *APOE _Ɛ_4* (Missing = 6) | |
| *CAIDE* (Missing = 10) | |
| Age | BMI |
| Sex | Cholesterol |
| Years of education (Missing =3) | Physical activity |
| Systolic blood pressure | APOE _Ɛ_4 (Missing = 6) |
| **Neuropsychological assessments** (Missing = 30) | |
| *COGNITO* (Missing = 1) | |
| Working memory (the simultaneous presentation of auditory and visual attention tasks assessed by subtracting the time taken in milliseconds on this double task from the visual task alone)  The score range was -14620 – 8967, a higher score indicated poorer performance. | Working memory (the simultaneous presentation of auditory and visual attention tasks assessed by total number of correct answers for visual form recognition on this double task)  The score range was -1 – 10, a higher score indicated better performance. |
| Narrative recall (total number of correct elements on immediate recall of a story with a temporal progression requiring attention to macrostructure)  The score range was 0 – 26, a higher score indicated better performance. | Description recall (total number of correct elements recalled of a description without thematic progression requiring attention to microstructure and recall of spatial location. The narrative and description recall are similar in terms of word frequency in the language and syntactic structure)  The score range was 0 – 27, a higher score indicated better performance. |
| Implicit memory (difference in the number of steps in the progressive build-up of names on the screen required for recognition between names never seen and number of names previously learnt in an immediate recall task)  The score range was -0.8 – 3.4, a higher score indicated better performance. | Name-face association (number of faces recognized after a delay from a series of 18 faces of which 9 have been previously shown with their corresponding names)  The score range was 0 – 9, a higher score indicated better performance. |
| Form perception: number of correct answers in the matching of complex forms to a multiple-choice array  The score range was 2 – 8, a higher score indicated better performance. | Form perception speed: mean time taken in milliseconds for each trial  The score range was 2486 – 12042, a higher score indicated poorer performance. |
| Phoneme comprehension (number of correct responses in the matching of a word with an image presented as part of a multiple-choice array including semantic, morphological and phonetic distractors)  The score range was 6 – 9, a higher score indicated better performance. | Phoneme comprehension speed (mean time taken in milliseconds to perform)  The score range was 818 – 2769, a higher score indicated poorer performance. |
| Verbal fluency (total sum of the number of words generated in 30s using both a semantic (vegetables) and phonemic (letter P) cue)  The score range is 1 – 47, a higher score indicates better performance. |  |
| *VSTMBT* (Missing = 29) | |
| Shape only condition (the percentage of correctly recognized items)  The score range was -0.9375 – 1.00, a higher score indicated better performance.  Shape-colour binding condition (the percentage of correctly recognized items)  The score range was -0.625 – 1.00, a higher score indicated better performance. | |
| **Lifestyle activities** | |
| *Lifetime of Experiences Questionnaire* | |

Note: APOE _Ɛ_4, Apolipoprotein _Ɛ_4; CAIDE, Cardiovascular Risk Factors, Aging and Dementia; BMI, body mass index; VSTMBT, Visual Short-Term Memory Binding task.

| Supplementary Table 2. Description of cognitive tasks and measures | | | |
| --- | --- | --- | --- |
| Cognitive Tasks | Measures | Task Description | Descriptive Statistics (N = 461) |
| Working memory | Total number of correct answers | Dual task: The subject must locate the targeted shapes and count the sounds. | Mean = 9.84 SD = 0.64 Range = 11 Missingness = 1 |
|  | Mean time (ms) difference in milliseconds between the dual task and a simple form recognition task | Dual task: The subject must locate the targeted shapes and count the sounds.  Simple task: Subject must locate the targeted shapes only. | Mean = -162.60 SD = 2962.26 Range = 16518  Missingness = 0 |
| Narrative recall | Total number of correct answers | The subject must recall a series of elements which have a logical sequence (a short story). | Mean = 14.40 SD = 4.16 Range = 26 Missingness = 0 |
| Description recall | Total number of correct answers | The subject must recall a series of elements which have a visual sequence (a short description). | Mean = 13.76 SD = 4.41 Range = 27 Missingness = 0 |
| Implicit memory | Difference between the number of names never seen and the number of names already learned | The subject must recognize as soon as possible a name which is constructed progressively on the screen. | Mean = 1.02 SD = 0.59 Range = 4.2 Missingness = 0 |
| Name-face association | The number of correctly recognized faces and their corresponding names | The subject must decide whether a face on the screen appeared before and if yes, what the person's name is. | Mean = 5.16 SD = 2.08 Range = 9 Missingness = 0 |

| Supplementary Table 2. Description of cognitive tasks and measures (continued) | | | |
| --- | --- | --- | --- |
| Form matching | Total number of correct answers | The subject must discriminate form and line orientation by matching a sample complex figure to one of six figures. Distractor figures are designed to detect visuospatial field neglect and difficulties with line orientation. | Mean = 6.32 SD = 1.18 Range = 6 Missingness = 0 |
|  | Mean time (ms) for correct answers |  | Mean = 5735.89 SD = 1538.98 Range = 9556 Missingness = 0 |
| Phoneme comprehension | Total number of correct answers | The subject must choose an object illustrating a presented word among 6 objects which include shape, phonetic and semantic distractors. | Mean = 8.61 SD = 0.61 Range = 3 Missingness = 0 |
|  | Mean time (ms) for correct answers |  | Mean = 1410.36 SD = 249.17 Range = 1951 Missingness = 0 |
| Verbal fluency | Total number of correct answers | The subject must name all the words they can think of within one minute based on semantic and phonetic cues. | Mean = 27.29 SD = 6.69 Range = 46 Missingness = 0 |
| VSTMBT-shape only | Total number of correct answers | The subject must recall stimuli that were shapes after a short period of retention. | Mean = 0.85 SD = 0.18 Range = 1.94 Missingness = 29 |
| VSTMBT-shape colour binding | Total number of correct answers | The subject must recall stimuli that were combinations of shapes and colours after a short period of retention. | Mean = 0.47 SD = 0.24 Range = 1.63 Missingness = 29 |
| Key: VSTMBT, visual short-term memory binding test; SD, standard deviation | | | |

Supplementary Table 3a. APOE – Regression coefficients for Episodic and Relational Memory (Component 1) including interaction terms

| Model summary | | R^2^ | F | p |
| --- | --- | --- | --- | --- |
|  |  | 0.11 | 11.10 | <0.001 |
| DV | IV | β (SE) | | p |
| Component 1 | APOE _Ɛ_4 | -0.49 (0.31) | | 0.11 |
|  | APOE _Ɛ_4*Sex | 0.35 (0.18) | | 0.06 |
|  | Age | -0.02 (0.01) | | 0.07 |
|  | Sex | 0.38 (0.12) | | 0.001 |
|  | Years of education | 0.04 (0.01) | | 0.003 |

Note: unstandardized coefficients β and standard error (SE) were reported. DV, dependent variable; IV, independent variable; APOE _Ɛ_4, Apolipoprotein _Ɛ_4.

Supplementary Table 3b. CAIDE (excl. APOE status) – Regression coefficients for Episodic and Relational Memory (Component 1)

| Model summary | | R^2^ | F | p |
| --- | --- | --- | --- | --- |
|  |  | 0.02 | 10.15 | 0.002 |
| DV | IV | β (SE) | | p |
| Component 1 | CAIDE  (excl. APOE status) | -0.08 (0.02) | | 0.002 |

Note: unstandardized coefficients β and standard error (SE) were reported. DV, dependent variable; IV, independent variable; CAIDE, Cardiovascular Risk Factors, Aging and Dementia; APOE, Apolipoprotein.

Supplementary Table 3c. CAIDE – Regression coefficients for Episodic and Relational Memory (Component 1) on West London site (208 participants)

| Model summary | | R^2^ | F | p |
| --- | --- | --- | --- | --- |
|  |  | 0.02 | 3.68 | 0.057 |
| DV | IV | β (SE) | | p |
| Component 1 | CAIDE | -0.05 (0.03) | | 0.057 |

Note: unstandardized coefficients β and standard error (SE) were reported. DV, dependent variable; IV, independent variable; CAIDE, Cardiovascular Risk Factors, Aging and Dementia.

Supplementary Table 3d. CAIDE – Regression coefficients for Episodic and Relational Memory (Component 1) including interaction terms

| Model summary | | R^2^ | F | p |
| --- | --- | --- | --- | --- |
|  |  | 0.02 | 5.41 | 0.01 |
| DV | IV | β (SE) | | p |
| Component 1 | CAIDE | -0.11 (0.05) | | 0.03 |
|  | CAIDE*Sex | 0.04 (0.03) | | 0.21 |

Note: unstandardized coefficients β and standard error (SE) were reported. DV, dependent variable; IV, independent variable; CAIDE, Cardiovascular Risk Factors, Aging and Dementia.

Supplementary Table 3e. FHD – Regression coefficients for Episodic and Relational Memory (Component 1)

| Model summary | | R^2^ | F | p |
| --- | --- | --- | --- | --- |
|  |  | 0.10 | 12.94 | <0.001 |
| DV | IV | β (SE) | | p |
| Component 1 | FHD | 0.08 (0.09) | | 0.38 |
|  | Age | -0.02 (0.01) | | 0.05 |
|  | Sex | 0.51 (0.09) | | <0.001 |
|  | Years of education | 0.04 (0.01) | | 0.003 |

Note: unstandardized coefficients β and standard error (SE) were reported. DV, dependent variable; IV, independent variable; FHD, family history of dementia.

Supplementary Table 4a. APOE – Regression coefficients for Multisensory Processing (Component 2)

| Model summary | | R^2^ | F | p |
| --- | --- | --- | --- | --- |
|  |  | 0.006 | 0.68 | 0.61 |
| DV | IV | β (SE) | | p |
| Component 2 | APOE _Ɛ_4 | -0.13 (0.10) | | 0.18 |
|  | Age | 0.01 (0.01) | | 0.50 |
|  | Sex | -0.01 (0.10) | | 0.92 |
|  | Years of education | 0.01 (0.02) | | 0.56 |

Note: unstandardized coefficients β and standard error (SE) were reported. DV, dependent variable; IV, independent variable; APOE _Ɛ_4, Apolipoprotein _Ɛ_4.

Supplementary Table 4b. CAIDE – Regression coefficients for Multisensory Processing (Component 2)

| Model summary | | R^2^ | F | p |
| --- | --- | --- | --- | --- |
|  |  | 0.002 | 1.05 | 0.31 |
| DV | IV | β (SE) | | p |
| Component 2 | CAIDE | -0.02 (0.02) | | 0.31 |

Note: unstandardized coefficients β and standard error (SE) were reported. DV, dependent variable; IV, independent variable; CAIDE, Cardiovascular Risk Factors, Aging and Dementia.

Supplementary Table 4c. FHD – Regression coefficients for Multisensory Processing (Component 2)

| Model summary | | R^2^ | F | p |
| --- | --- | --- | --- | --- |
|  |  | 0.003 | 0.36 | 0.84 |
| DV | IV | β (SE) | | p |
| Component 2 | FHD | 0.07 (0.10) | | 0.46 |
|  | Age | 0.01 (0.01) | | 0.47 |
|  | Sex | -0.02 (0.10) | | 0.87 |
|  | Years of education | 0.01 (0.02) | | 0.53 |

Note: unstandardized coefficients β and standard error (SE) were reported. DV, dependent variable; IV, independent variable; FHD, family history of dementia.

Supplementary Table 5a. APOE – Regression coefficients for Short-term Memory Binding (Component 3)

| Model summary | | R^2^ | F | p |
| --- | --- | --- | --- | --- |
|  |  | 0.03 | 2.88 | 0.02 |
| DV | IV | β (SE) | | p |
| Component 3 | APOE _Ɛ_4 | 0.02 (0.10) | | 0.87 |
|  | Age | -0.02 (0.01) | | 0.05 |
|  | Sex | -0.17 (0.10) | | 0.07 |
|  | Years of education | 0.03 (0.02) | | 0.03 |

Note: unstandardized coefficients β and standard error (SE) were reported. DV, dependent variable; IV, independent variable; APOE _Ɛ_4, Apolipoprotein _Ɛ_4.

Supplementary Table 5b. CAIDE – Regression coefficients for Short-term Memory Binding (Component 3)

| Model summary | | R^2^ | F | p |
| --- | --- | --- | --- | --- |
|  |  | 0.007 | 3.13 | 0.08 |
| DV | IV | β (SE) | | p |
| Component 3 | CAIDE | -0.03 (0.02) | | 0.08 |

Note: unstandardized coefficients β and standard error (SE) were reported. DV, dependent variable; IV, independent variable; CAIDE, Cardiovascular Risk Factors, Aging and Dementia.

Supplementary Table 5c. FHD – Regression coefficients for Short-term Memory Binding (Component 3)

| Model summary | | R^2^ | F | p |
| --- | --- | --- | --- | --- |
|  |  | 0.03 | 2.90 | 0.02 |
| DV | IV | β (SE) | | p |
| Component 3 | FHD | -0.03 (0.10) | | 0.74 |
|  | Age | -0.02 (0.01) | | 0.06 |
|  | Sex | -0.17 (0.10) | | 0.08 |
|  | Years of education | 0.03 (0.02) | | 0.04 |

Note: unstandardized coefficients β and standard error (SE) were reported. DV, dependent variable; IV, independent variable; FHD, family history of dementia.

Supplementary Table 6a. Lifestyle & Sex – Regression coefficients for Episodic and Relational Memory (Component 1) including interaction terms in APOE _Ɛ_4- group

| Model summary | | *R^2^* | *F* | *p* |
| --- | --- | --- | --- | --- |
|  |  | 0.12 | 5.33 | <0.001 |
| DV | IV | *β (se)* | | *p* |
| Component 1 | Specific | 0.01 (0.04) | | 0.82 |
|  | Non-specific | 0.01 (0.05) | | 0.83 |
|  | Sex*Specific | 0.01 (0.02) | | 0.79 |
|  | Sex*Non-specific | 0.03 (0.03) | | 0.36 |
|  | Age | -0.03 (0.01) | | 0.02 |
|  | Sex | 0.36 (0.12) | | 0.003 |
|  | Years of education | 0.03 (0.02) | | 0.07 |

Note: unstandardized coefficients β and standard error (SE) were reported. DV, dependent variable; IV, independent variable; APOE _Ɛ_4-, Apolipoprotein _Ɛ_4 genotype negative.

Supplementary Table 6b. Lifestyle & Sex – Regression coefficients for Episodic and Relational Memory (Component 1) including interaction terms in FHD+ group

| Model summary | | R^2^ | F | p |
| --- | --- | --- | --- | --- |
|  |  | 0.21 | 8.63 | <0.001 |
| DV | IV | β (SE) | | p |
| Component 1 | Specific | -0.02 (0.05) | | 0.65 |
|  | Non-specific | -0.03 (0.07) | | 0.68 |
|  | Sex*Specific | 0.03 (0.03) | | 0.35 |
|  | Sex*Non-specific | 0.05 (0.04) | | 0.21 |
|  | Age | -0.04 (0.01) | | 0.004 |
|  | Sex | 0.67 (0.14) | | <0.001 |
|  | Years of education | 0.03 (0.02) | | 0.16 |

Note: unstandardized coefficients β and standard error (SE) were reported. DV, dependent variable; IV, independent variable; FHD+, positive family history of dementia.

Supplementary Table 6c. Lifestyle & Sex – Regression coefficients for Episodic and Relational Memory (Component 1) including interaction terms in FHD- group

| Model summary | | R^2^ | F | p |
| --- | --- | --- | --- | --- |
|  |  | 0.11 | 3.68 | <0.001 |
| DV | IV | β (SE) | | p |
| Component 1 | Specific | -0.06 (0.04) | | 0.16 |
|  | Non-specific | 0.07 (0.05) | | 0.16 |
|  | Sex*Specific | 0.04 (0.03) | | 0.14 |
|  | Sex*Non-specific | -0.01 (0.03) | | 0.68 |
|  | Age | -0.003 (0.01) | | 0.78 |
|  | Sex | 0.31 (0.13) | | 0.01 |
|  | Years of education | 0.03 (0.02) | | 0.13 |

Note: unstandardized coefficients β and standard error (SE) were reported. DV, dependent variable; IV, independent variable; FHD-, negative family history of dementia.

Supplementary Table 6d. Lifestyle & Sex – Regression coefficients for Episodic and Relational Memory (Component 1) including interaction terms in high CAIDE group

| Model summary | | R^2^ | F | p |
| --- | --- | --- | --- | --- |
|  |  | 0.22 | 7.20 | <0.001 |
| DV | IV | β (SE) | | p |
| Component 1 | Specific | -0.03 (0.05) | | 0.57 |
|  | Non-specific | 0.13 (0.06) | | 0.053 |
|  | Sex*Specific | 0.04 (0.03) | | 0.22 |
|  | Sex*Non-specific | -0.04 (0.04) | | 0.33 |
|  | Age | -0.02 (0.02) | | 0.42 |
|  | Sex | 0.64 (0.15) | | <0.001 |
|  | Years of education | 0.04 (0.02) | | 0.13 |

Note: unstandardized coefficients β and standard error (SE) were reported. DV, dependent variable; IV, independent variable; CAIDE, Cardiovascular Risk Factors, Aging and Dementia.

Supplementary Table 6e. Lifestyle & Sex – Regression coefficients for Episodic and Relational Memory (Component 1) including interaction terms in low CAIDE group

| Model summary | | R^2^ | F | p |
| --- | --- | --- | --- | --- |
|  |  | 0.11 | 4.53 | <0.001 |
| DV | IV | β (SE) | | p |
| Component 1 | Specific | -0.08 (0.05) | | 0.08 |
|  | Non-specific | -0.05 (0.06) | | 0.40 |
|  | Sex*Specific | 0.05 (0.03) | | 0.07 |
|  | Sex*Non-specific | 0.06 (0.03) | | 0.10 |
|  | Age | -0.02 (0.01) | | 0.15 |
|  | Sex | 0.41 (0.12) | | 0.001 |
|  | Years of education | 0.03 (0.02) | | 0.12 |

Note: unstandardized coefficients β and standard error (SE) were reported. DV, dependent variable; IV, independent variable; CAIDE, Cardiovascular Risk Factors, Aging and Dementia.
